# CORRELATION BETWEEN MEASURED PARAMETERS OF RISK AND PROGNOSIS IN SUBJECTS WITH CHRONIC PERIODONTITIS

**DOI:** 10.1101/2022.10.17.22281156

**Authors:** Rampalli Viswa Chandra, Tulasi Shravan

## Abstract

**Aims:** In most ofcases, clinical parameters associated with poor prognosis are the same as the factors associated with increased risk, but the relationship between risk and prognosis remains unclear.The objective of this study was to evaluate the correlation between measured parameters of predictive risk and prognosisin subjects with chronic periodontitis.

**Methods:** 300 subjects participated in the study. Modified periodontal risk assessment model (MPRA) was used to assess the risk and prognosis was evaluated by using McGuire and Nunn prognostic criteria.

**Results:** Among the subjects, 57.3%, 38% and 4.7% were categorized as having high, low and medium risk respectively. Assessment of prognosis among study subjects showed that 38.0%, 24.0%, 17.7%, 17.0%, 2.0%and1.3% had good, fair, excellent, poor, hopeless and questionable prognosis respectively. Though majority of the subjects had good prognosis (114 subjects;38%), there was a substantial variability in the distribution of the measured parameters as per the risk scores within this cohort. A strong positive correlation was seen between prognosis and probing depth (PD)≥5mm. There was a weak, but statistically significant correlation between predictive risk from MPRA and various types of prognosis(r_s_=0.507;p<0.001).

**Conclusion:** The measures used to assess risk and prognosis are almost similar;butthe weak correlation between risk and prognosisseems to suggest that an increased risk of developing periodontal disease need notnecessarily indicate a bad prognosis.

## INTRODUCTION

Chronic periodontitis is an inflammatory disease that affects 8% to 10% of population.^1^ The host response to various etiologic agents vary among patients with periodontitis; hence it is clinically important to determine the various factors that increase a person’s chances of developing periodontal disease. The identification of these periodontal risk factors have contributed greatly towards understanding the pathogenesis of periodontitis.^2^ A more detailed and comprehensive assessment of a patient’s risk characteristics would be essential to calculate more accurately their individual risk and to formulate prognosis and make informed treatment decisions.^2,3^ Risk of periodontal disease is significantly related to oral health related quality of life and one method of identifying and analyzing potential risk factors is by using a risk assessment model.^2-5,6,7^ Various risk assessment models are in vogue and one such model is themodified periodontal risk assessment model (MPRA).^5^ This modified PRA model is easy to generate and obtain, uses retrospective and current data to assess the risk of periodontal disease in contrast to the original assessment model which is more complicated to use.

Prognosis is derived by those characteristics that may influence the outcome once the disease is established.^8,9^ Traditionally based on tooth mortality,^8-11^ prognosis establishes the range of disease consequences and prognostic factors are clinical factors that are significant in predicting tooth loss as a result of periodontal disease.^8^ Periodontal prognosis is based on clearly defined objective criteria and the essential element of prognosis is the timing of the projection.^8-11^ Periodontal prognostication is dynamic, thus it should be reevaluated periodically as treatment and maintenance progress.^11^

In most of the cases,clinical parameters associated with poor prognosis are same as the factors associated with increased risk, but the relationship between risk and prognosis still remains unclear.^10,12-14^ Findings from the Piedmont 65+ dental study^12^ seem to indicate that risk factors are important in explaining disease progression;however, parameters such as probing depth (PD), furcation, mobility, bone loss and smoking which area part of the diagnostic criteria of various risk assessment models^2,3,5,7,9^ are also used in the evaluation of prognosisas well.^8,10,12^ With research mainly focussing on risk factors^12,13^ and with relatively fewer prognostic factors,^8,10^ the numerical disparity between the number of identifiable risk and prognostic factors also seems to have contributed to the general lack of consensus on the relationship between risk and prognosis.^15-20^

Thoughrisk and prognosis represent the onset of disease *vs*the consequences of a disease,^7,9,12^ understanding this relationship in a subject with periodontitis is crucial because of the following reasons; 1. The effect of identified risk factors on long-term prognosis can be evaluated^3,5,6^ 2. Identifying specific risk factors that have maximum impact on eventualprognosis would be of immense aid to a clinician enabling him to customize treatment options based on eradicating the risk factors.^19,21,22^ 3. Identifying which clinical parameters best determine risk and prognosiscan also alert a clinician to be on a lookout for these signs in a subject with developing or established periodontitis.^2,3,8,10,11^ In light of the expected benefits, the objective of this study was toevaluate the correlation between measured parameters ofpredictive risk and prognosisin subjects with chronic periodontitis.

## MATERIALS AND METHODS

### Study design and source of data

A sample of 300(57.3% males; mean age: 48.67±18.22 years) subjects with chronic periodontitis were randomly selected from the patient pool of subjects presenting to the Department of Periodontics and Implantology, for initial evaluation between April to September 2017. Inclusion criteria were subjects>30yrs diagnosed with chronic generalised Periodontitis based on gingival index score of >/=1 and attachment loss of >/=3mm in more than 30% of the sites with presence of atleast 20 natural teeth reporting for intial outpatient evaluation.

Subjects who underwent or were under periodontal therapy were excluded from the study.All cases were examined by threecalibratedperiodontists (DAT, PAK and AA) throughstandardized probing, automated charting and radiographic examination.The study was conducted in compliance with the institutional ethical committee’s charter and guidelines (Ref.no. SVSIDS/PERIO/3/2015) and informed consent was taken from all thesubjects.

#### Sample size

A sample size of 300 was calculated for an effect size of 0.16, with probability of α error being 0.05 and a power of 0.80.

### Assessment of Predictive Risk and Prognosis

All the participating subjects were assessed for various risk factors of periodontal disease by using a previously described model (modified periodontal risk assessment model/MPRA).^5^ Briefly, 8 measured parameters were evaluated: 1, percentage of sites with bleeding on probing (BOP); 2, number of sites with probing depth (PD) ≥5 mm; 3, number of teeth lost; 4, attachment loss (AL)/age ratio; 5, diabetic status; 6, smoking; 7, dental status – systemic factors interplay (DS-SFI); and 8, other background characteristics. BOP, PD, tooth loss and AL/age ratio measure the cumulative periodontal status*(Figure 1)*. Diabetic status and smoking are the risk factors, and stress and socio-economic factors are the risk determinants assessed in this model. The parameters were plotted on the radar chart which then generates a radar chart and calculates risk automatically*(Figure 2)*. All the measured parameters were assessedon a 0-5 risk scale andthe overall predictive risk was calculated as follows; A low-periodontal-risk patient has all the parameters in the low-risk area, or at the most two parameters in the moderate and high-risk area.A moderate-periodontal-risk patient has at least three parameters in the moderate-risk area and not more than one parameter in the high-risk area.A high-periodontal-risk patient has at least two parameters in the high-risk category.

The prognosis was evaluated by using McGuire and Nunn^8,10^ prognostic criteria, in which the prognosis was classified into excellent, good, fair, poor, questionable and hopeless prognosis as follows; Excellent prognosis: No bone loss, excellent gingival condition, good patient cooperation, no systemic or environmental factors.Good prognosis (one or more of the following): Control of the etiologic factors and adequate periodontal support as measured clinically and radiographically to assure the tooth would be relatively easy to maintain by the patient and clinician assuming proper maintenance.Fair Prognosis (one or more of the following): Approximately 25% attachment loss as measured clinically and radiographically and/or class I furcation involvement. The location and depth of the furcations would allow proper maintenance with good patient compliance.Poor Prognosis (one or more of the following): 50% attachment loss with class II furcations. The location and depth of the furcations would allow proper maintenance, but with difficulty.Questionable Prognosis (one or more of the following): Greater than 50% attachment loss resulting in a poor crown-to-root ratio. Poor root form, class II furcations not easily accessible to maintenance care or class III furcations, 2+ mobility or greater, significant root proximity.Hopeless Prognosis: Inadequate attachment to maintain the tooth. Extraction performed or suggested.

### Correlation between measured parameters of risk and prognosis

Extraction of relevantdata was done by fourcalibrated examiners (SN, JN, PAK and DAT). Predictive risk and Prognosis were assessed by a different set of calibrated examiners (AAR and RVC).

#### Statistical Analysis

Data was analysed by using SPSS ver.21^®^ statistical software package. Descriptive statistics and frequency distribution were analysed. Spearman’s Rho correlation test was used to test the correlation between prognosis and MPRA and the other variables. p≤0.05 was considered statistically significant.

## RESULTS

The objective of this study was to evaluate the correlation between measured parameters of predictive risk and prognosisin 300 subjects with chronic periodontitis. Parameters were graded according a pre-set scale and the risk assessment polygon was generated based on the data assessed *(Figure 1,2)*.

*Table 1* shows the distribution of the constituent parameters as per the risk scores and their association with various types of prognosis. Though majority of the subjects had a good prognosis (114 subjects;38%), there was a substantial variability in the distribution of the measured variables as per the risk scoreswithin the cohort.A maximum risk assessment score of 5 was seen for parameters BOP (65 subjects; 57%), PD ≥ 5mm (48 subjects; 42.1%) and background characteristics (52 subjects; 45.6%). A risk assessment score of 0 was seen for parameters tooth loss (84 subjects; 73.7%), smoking (101 subjects; 88.6%)anddiabetic status (92 subjects; 80.7%). A risk assessment score of 1 and 2 was seen for parameters attachment loss /age ratio (99 subjects; 86.8%) and dental status-systemic factor interplay (103 subjects; 90.4%) respectively. However, within all groups, subjects with excellent prognosis largely had 5-9% of sites with BOP (n=16;30.2%), 1-2 sites with PD ≥ 5mm(n=20;37.7%), no tooth loss (n=47;88.l7%), were non-smokers (n=52;98.1%), hadAL/age ratio≤ 0.25 (n=1;1.9%), were non-diabetic subjects (n=47;88.7%) and had the DS-SFI score of 2 (n=48;90.6%)

**Table 1:**
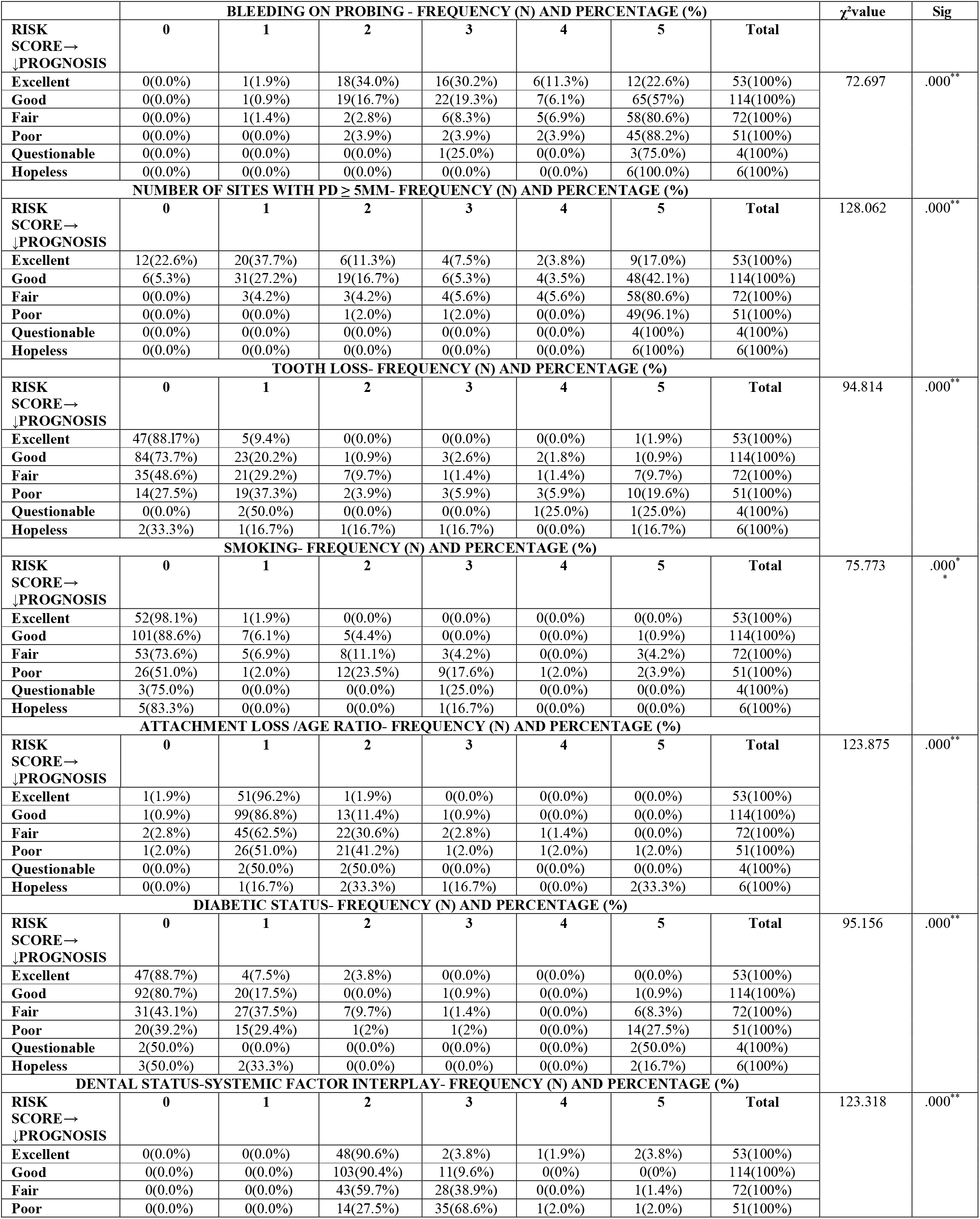
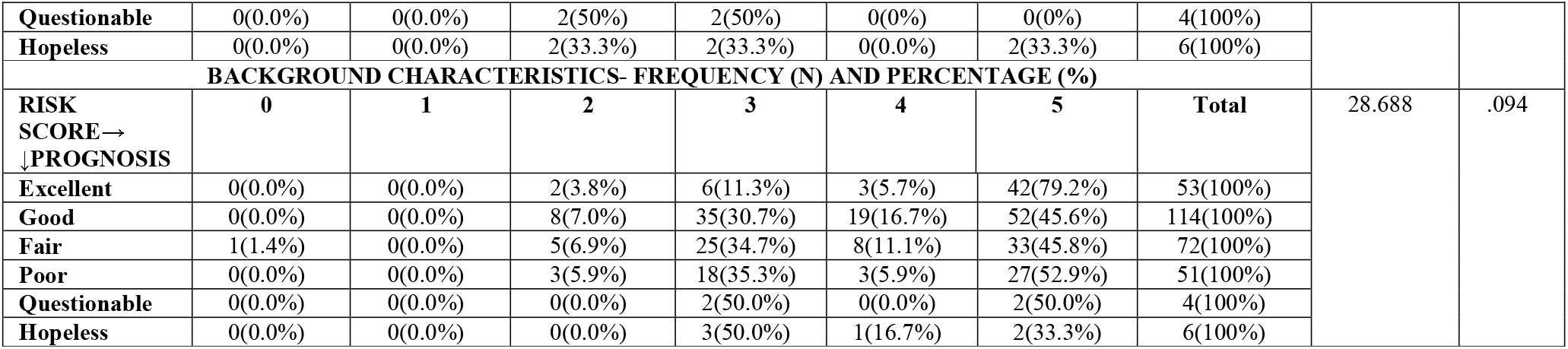
Frequency distribution of the constituent parameters as per the risk scores and their association with prognosis.

*Table 2* shows the frequency distribution of the measures of predictive risk and types of prognosis among the study subjects. Among the subjects, 57.3%, 38% and 4.7% were categorized as having high, low and medium risk respectively. The distribution of these variables in the study population were statistically significant*(p* ≤ *0*.*001)*. Assessment of prognosis among study subjects showed that 38.0% had good prognosis, 24.0% had fair prognosis, 17.7% had excellent prognosis, 17.0% had poor prognosis, 2.0% had hopeless prognosis and only1.3% had questionable prognosis. The distribution of these variables in the study population were statistically highly significant *(p* ≤ *0*.*001)*.

**Table 2:** Frequency distribution of the measures of predictive risk and types of prognosis among the study subjects

| *Variable* | *Category/Type* | *n* | *%* | *χ<sup>2</sup>* | *p-value* |
| --- | --- | --- | --- | --- | --- |
| **MPRA**<br>**Assessment** | Low | 114 | 38.0 | 127.760 | 0.000** |
|  | Medium | 14 | 4.7 |  |  |
|  | High | 172 | 57.3 |  |  |
|  | **Total** | 300 | 100.0 |  |  |
| **Prognosis** | Excellent | 53 | 17.7 | 172.840 | 0.000** |
|  | Good | 114 | 38.0 |  |  |
|  | Fair | 72 | 24.0 |  |  |
|  | Poor | 51 | 17.0 |  |  |
|  | Questionable | 4 | 1.3 |  |  |
|  | Hopeless | 6 | 2.0 |  |  |
|  | **Total** | 300 | 100.0 |  |  |

*Table 3* shows the correlations between prognosis and the constituent parameters of MPRA. No correlation was seen between prognosis and background characteristics. Weak positive correlation was seen between prognosis and smoking, moderate positive correlation was seen between prognosis and BOP, tooth loss, attachment loss / age ratio, diabetes, DS-SFI and tooth loss and a strong positive correlation was seen between prognosis and PD≥5mm.

**Table 3:** Correlations between prognosis and the constituent parameters of MPRA.

|  | *Prognosis* | *Smoking* | *BOP* | *PD ≥ 5 mm* | *Tooth Loss* | *AI/Age Ratio* | *Diabetes* | *Dental Status-Systemic Factors Interplay* | *Background characteristics* |
| --- | --- | --- | --- | --- | --- | --- | --- | --- | --- |
| **Prognosis** | - | .370** | .461*** | .603**** | .466*** | .400*** | .431*** | .482*** | -.158<sup>+</sup> |

*Spearman's Rho Correlation* \* Very weak positive correlation, \*\* weak positive correlation, \*\*\* moderate positive correlation, \*\*\*\*strong positive correlation, <sup>+</sup> statistically not significant

*Table 4* shows thecorrelations between the predictive risk from MPRA and various types of prognosis. There was a weak, but statistically significant correlation between these two variables. The following trends were also observed in these correlations. 50.9% and 34.2% of subjects whose predictive risk was low had good and excellent prognosis respectively.35.7% and 28.6% of subjects with predictive risk categorized as moderate had good or fair prognosis and 32% and 26.2% of subjects with high predictive riskhad fair and poor prognosis respectively.

**Table 4:** Correlation* between the predictive risk from MPRA and various types of prognosis.

|  | **Excellent** | **Good** | **Fair** | **Poor** | **Questionable** | **Hopeless** |
| --- | --- | --- | --- | --- | --- | --- |
| **Low** | 39<br>(34.2%) | 58<br>(50.9%) | 13<br>(11.4%) | 4<br>(3.9%) | 0 | 0 |
| **Moderate** | 3<br>(21.4%) | 5<br>(35.7%) | 4<br>(28.6%) | 2<br>(14.3%) | 0 | 0 |
| **High** | 11<br>(6.4%) | 51<br>(29.7%) | 55<br>(32%) | 45<br>(26.2%) | 4<br>(2.3%) | 6<br>(3.5%) |

\*Spearman's Correlation ( $r_s=0.507; p<0.001$ )

## DISCUSSION

The primary objective of this study was to evaluate the correlation between predictive risk and prognosisin subjects with chronic periodontitis. Majority (38%) of the subjects had a good prognosis; however, within this cohort itself, a maximum risk assessment score of 5 was seen for parameters BOP (65 subjects with >25% of sites with BOP), PD ≥ 5mm (48 subjects with >9 sites with PD≥5mm)and background characteristics (52 subjects who were umemployed/living in a very stressful environment). Claffey et al.,^15^ and Badersten et al.,^16^ showed that BOP between 20% and 30% indicates a higher risk for disease progression and studies of Grbic et al.,^11^ and McGuire et al.,^8^ showed that deep probing depths and attachment loss are associated with future periodontal breakdown.The numerical superiority of this cohort might have contributed to this finding which seems to go against the expected norm of healthy sites having better prognosis. Subjects with excellent prognosis largely demonstrated findings associated with excellent periodontal health; 1-2 sites with PD ≥ 5mm, no tooth loss, were non-smokers and non-diabetics, hadAL/age ratio≤ 0.25and had the DS-SFI score of 2. Similar trend was seen for background characteristics as well with subjects with poor(52.9%), questionable (50%) and hopeless prognosis (33.3%) showing higher risk scores than subjects with good prognosis (45.6%).

There was a significant distribution of the measures of predictive risk and types of prognosis among the study subjects. A large percentage of subjects had risk and prognosis categorized as having high (57.3%) and good (38%)respectively. When correlations between prognosis and the constituent parameters of MPRA were analysed, moderate to strong positive correlations were seen between prognosis and BOP, tooth loss, attachment loss / age ratio, diabetes, dental status-systemic factor interplayand tooth loss. When regression analysis was performed with prognosis as the dependent variable and constituent parameters of MPRA as the predictors. The same variables except BOP (PD ≥ 5mm, tooth loss, smoking, AL/age ratio, diabetes and DS-SFI), predicted their influence on the dependent variable (prognosis) significantly well. The impact of these parameters on prognosis have been established conclusively through a plethora of studies. Periodontitis results in an increased susceptibility to tooth loss^18-20^ and tooth lossis a better indicator than probing as a marker of lifetime oral health and is less prone to measurement error.^19^ A wealth of data has established the relationship between smoking and periodontitis and cigarette smoking affects the periodontium at many levels.^21-27^ AL/age ratiois related to the age and as people age, their risk for developing periodontitis increases.^1,21,28,29^ Neely et al.,^28^ observed that age remains as a significant risk factor for progression of mean attachment loss after adjusting for important co-factors in a 20-yearfollow up analysis of the natural history of periodontal disease. The risk of having periodontitisis also known to increase two-fold in 50+ years-old subjects when compared with younger subjects after adjusting for important co-factors.^28,29^ There is ample evidence linking diabetes mellitus and periodontitis.^21,30,31^ In our study 70.7% had other dental and systemic problems affecting the periodontium (as pertaining to DS-SFI). Studies have demonstrated significant increase in probing pocket depths, attachment loss and gingival inflammation when certain dental and systemic conditions conditions having a potential to disrupt periodontal homeostasis co-exist.^32-39^

52.7% of the subjects in our study were unemployed, a potentially stressful situation, and subjects with excellent prognosis(34.0%) had BOP in 5-9% of total sites.BOP and background characteristics did not have a significant effect on prognosis in the regression model.The possible relationship between periodontal disease and socioeconomic status and stressremains unproven and is subject to debate.^21,40,41,42^ Haas et al.,^21^ showed that periodontal attachment loss was not statistically associated with skin color and socio-economic status in Brazilian population and Hugoson et al.,^42^ stated that individuals with the ability to cope withstressful stimulipresent with improved measures of periodontal disease. Stress and socioeconomic factors may show different outcomes of periodontal disease depending on the interplay between the microbiota, host response and stress trait of an individual and a dominance of other factors such as PD, AL/age ratio, tooth loss and systemic factors over stress cannot be ruled out.^2,4-6,21,40,41,42^ BOP is useful for predicting the progression of periodontitis^43^ and BOP affecting 20% and 30%of sites is associated with increased risk and bad prognosis.^15,16^ BOP was associated with variable findings depending on the cohort in this study. Most of the subjects with excellent prognosis (34.0%) had BOP in 5-9% of the sites which is lessthan the 20-30% range to impact the prognosis of a subject. However, 57% of subjects with good prognosis showed >25% of sites with BOP. BOP is considered to be an important parameter to assess treatment response rather than to assess disease severity^1,2,4,5,7,12^ and this may have contributed to the contrasting pattern seen based on the type of prognosisand findings seem to challenge the emphasis placed on BOP as a prognostic indicator.

In our study, 74.7% had AL/age ratio≤ 0.25, 60.7% subjects did not have tooth loss (only 2.3% had 7-8 teeth lost), 80% were non-smokers, 65.0% had their fasting glucose blood sugar levels < 102 mg/dl, 70.7% had dental problems affecting periodontium.Assessment of correlation between MPRA scores and prognosis scores showed weakbut statistically significant correlation.Predominantly, subjectsin low risk group had good and excellent prognosis, while subjects in moderate risk group had good and fair prognosis and subjects in high risk group had fair and poor prognosis. Parameters that measure predictive risk also aid in identifying subjects with different rates and types of progressionand prognosis.^2,4^ The distinction between these two factors is vital as it is the prognosis and not the risk that eventually determines the treatment plan of a subject.^2,19,21,44^ Thus in this context, a lack of a strong direct relationship between predictive risk and prognosis may not be detrimental to the patient as assessment and control of common factors such as probing depth, tooth loss, smoking, attachment loss, diabetes and other dental problems can be used to effectively assess predictive riskand treat patients to improve outcomes.^2-6,12-14,44^

Like previous studies on the relationship between risk and prognosis, this study has limitations. One limitation is that this is a single-centre study.A multi-centre study with a larger-sized cohort may identify significant associations and account for unusual variations. We also sought to establish a correlation between measured parameters of risk and prognosis;butprognosis has its own set of measured parameters such as attachment loss, poor root form, furcation measurement,access to maintenance care, mobility and root proximity.^8-10^ However, this may not be a major limitation asdata gleaned fromcomprehensive periodontal examinationand measurement of parameters such as BOP, PD, tooth loss, smoking status, attachment loss/age ratio, diabetic status,DS-SFI and background characteristicsare similar to parameters used to establish periodontal prognosis.^4,5,10,13,44^

To conclude, the measures used to assess risk and prognosis are almost similar, butthe weak correlation between risk and prognosis seems to suggest that an increased risk of developing periodontal disease need notnecessarily indicate a bad prognosis.Parameters identified in this study are already an integral part ofexaminationprotocols for risk prediction and prognosis assessment at an individual level. Future studies should address the utility of these measures in formulating a customized treatment plan based on eradicating risk factors and improving prognosis.

## Data Availability

All data produced in the present study are available upon reasonable request to the authors

## REFERENCES

1. Loe H, Anerud A, Boysen H, Morrison E. Natural history of periodontal disease in man. Rapid, moderate and no loss of attachment in Sri Lankan laborers 14 to 46 years of age. J Clin Periodontol 1986;13:431–45.

2. Lang NP, Suvan JE, Tonetti MS. Risk factor assessment tools for the prevention of periodontitis progression a systematic review. J Clin Periodontol 2015;42:59–70.

3. Garcia RI, Compton R, Dietrich T. Risk assessment and periodontal prevention in primary care. Periodontol 2000 2016;71:10–21.

4. Lang NP, Tonetti MS. Periodontal risk assessment (PRA) for patients in supportive periodontal therapy (SPT). Oral Health Prev Dent 2003;1:7–16.

5. Chandra RV. Evaluation of a novel periodontal risk assessment model in patients presenting for dental care. Oral Health Prev Dent 2007;5:39–48.

6. Yang MC, Marks RG, Clark WB, Magnusson I. Predictive power of various models for longitudinal attachment level change. J Clin Periodontol 1992;19:77–83.

7. Nagarajan S, Chandra RV. Perception of oral health related quality of life among periodontal risk patients before and after periodontal therapy. Commun Dent Health 2012;29:90–4.

8. McGuire MK, Nunn ME. Prognosis versus actual outcome. II. The effectiveness of clinical parameters in developing an accurate prognosis. J Periodontol 1996;67:658–65.

9. Fan J, Nunn ME, Su X. Multivariate exponential survival trees and their application to tooth prognosis. Comput Stat Data Anal 2009;53:1110–21.

10. McGuire MK. Prognosis versus actual outcome: a long-term survey of 100 treated periodontal patients under maintenance care. J Periodontol 1991;62:51–8.

11. Grbic JT, Lamster IB, Celenti RS, Fine JB. Risk indicators for future clinical attachment loss in adult periodontitis. Patient variables. J Periodontol 1991;62:322–9.

12. Beck JD. Methods of assessing risk for periodontitis and developing multifactorial models. J Periodontol 1994;65:468–78.

13. Page RC, Martin JA, Loeb CF. The Oral Health Information Suite (OHIS): its use in the management of periodontal disease. J Dent Educ 2005;69:509–20.

14. American Academy of Periodontology. American Academy of Periodontology statement on risk assessment. J Periodontol 2008;79:202–9.

15. Claffey N, Nylund K, Kiger R, Garrett S, Egelberg J. Diagnostic predictability of scores of plaque, bleeding, suppuration, and probing pocket depths for probing attachment loss. 3 1/2 years of observation following initial therapy. J Clin Periodontol 1990;17:108–14.

16. Badersten A, Nilveus R, Egelberg J. Scores of plaque, bleeding suppuration and probing depth to predict probing attachment loss. J Clin Periodontol 1990;17:102–7.

17. Caffesse RG, Sweeney PL, Smith BA. Scaling and root planning with and without periodontal flap surgery. J Clin Periodontol 1986;13:205–10.

18. Kayser AF. Limited treatment goals -shortened dental arches. Periodontol 2000 1994;4:7–14.

19. Tu YK, Gilthorpe MS. Commentary: Is tooth loss good or bad for general health? Int J Epidemiol 2005;34:475–6.

20. Kwok V, Caton JG. Commentary: prognosis revisited: a system for assigning periodontal prognosis. J Periodontol 2007;78:2063–71.

21. Haas AN, Wagner MC, Oppermann RV, Rosing CK, Albandar JM, Susin C. Risk factors for the progression of periodontal attachment loss: a 5-year population-based study in south Brazil. J Clin Periodontol 2014;41:215–23.

22. Bergstrom J, Eliasson S. Cigarette smoking and alveolar bone height in subjects with a high standard of oral hygiene. J Clin Periodontol 1987;14:466–9.

23. Ismail AI, Burt BA, Eklund SA. Epidemiologic patterns of smoking and periodontal disease in the United States. J Am Dent Assoc 1983;106:617–21.

24. Van der Velden U, Varoufaki A, Hutter JW, et al. Effect of smoking and periodontal treatment on the subgingival microflora. J Clin Periodontol 2003;30:603–10.

25. Rawlinson A, Grummitt JM, Walsh TF, Ian Douglas CW. Interleukin 1 and receptor antagonist levels in gingival crevicular fluid in heavy smokers versus nonsmokers. J Clin Periodontol 2003;30:42–8.

26. Tipton DA, Dabbous MK. Effects of nicotine on proliferation and extracellular matrix production of human gingival fibroblasts in vitro. J Periodontol 1995;66:1056–64.

27. Kubota M, Tanno-Nakanishi M, Yamada S, Okuda K, Ishihara K. Effect of smoking on subgingival microflora of patients with periodontitis in Japan. BMC Oral Health 2011;11:1–6.

28. Neely AL, Holford TR, Loe H, Anerud A, Boysen H. The natural history of periodontal disease in man. Risk factors for progression of attachment loss in individuals receiving no oral health care. J Periodontol 2001;72:1006–15.

29. Albandar JM. Some predictors of radiographic alveolar bone height reduction over 6 years. J Periodontal Res 1990;25:186–92.

30. Emrich LJ, Shlossman M, Genco RJ. Periodontal disease in non-insulin dependent diabetes mellitus. J Clin Periodontol 1991;62:123–131.

31. Pucher J and Stewart J. Periodontal disease and diabetes mellitus. Curr Diab Rep 2004;4:46–50.

32. Kamil W, Al Habashneh R, Khader Y, Al Bayati L, Taani D. Effects of nonsurgical periodontal therapy on C-reactive protein and serum lipids in Jordanian adults with advanced periodontitis. J Periodontal Res 2011;46:616–21.

33. Loos BG. Systemic markers of inflammation in periodontitis. J Periodontol 2005;76:2106–15.

34. Grau AJ, Becher H, Ziegler CM, Lichy C, Buggle F, Kaiser C. Periodontal disease as a risk factor for ischemic stroke. Stroke 2004;35:496–501.

35. Kim HD, Sim SJ, Moon JY, Hong YC, Han DH. Association between periodontitis and hemorrhagic stroke among Koreans: a case–control study. J Periodontol 2010;81:658–65.

36. Persson GR, Persson RE. Cardiovascular disease and periodontitis: an update on the associations and risk. J Clin Periodontol 2008;35:362–79.

37. Andriankaja OM, Genco RJ, Dorn J, Dmochowski J, Hovey K, Falkner KL, Trevisan M. Periodontal disease and risk of myocardial infarction: the role of gender and smoking. Eur J Epidemiol 2007;22:699–705.

38. Niedzielska I, Janic T, Cierpka S, Swietochowska E. The effect of chronic periodontitis on the development of atherosclerosis: review of the literature. Med Sci Monit 2008;14:103–6.

39. Nishida M, Grossi SG, Dunford RG, Ho AW, Trevisan M, Genco RJ. Calcium and the risk for periodontal disease. J Periodontol 2000;71:1057–66.

40. Beck JD, Koch GG, Rozier RG, Tudor GE. Prevalence and risk indicators for periodontal attachment loss in a population of older community-dwelling blacks and whites. J Periodontol 1990;61:521–8.

41. Liu R, Desta T, He H, Graves DT. Diabetes alters the response to bacteria by enhancing fibroblast apoptosis. Endocrinology 2004;145:2997–3003.

42. Hugoson A, Ljungquist B, Breivik T. The relationship of some negative events and psychological factors to periodontal disease in an adult Swedish population 50 to 80 years of age. J Clin Periodontol 2002;29:247–53.

43. Sachan S. Risk Factor for Periodontitis. J Comput Eng 2014;16:88–92.

44. Lyle DM. Risk assessment a key to periodontal health promotion and disease prevention. Compend Contin Educ Dent 2014;35(6):392–396.

